# An *in silico* analysis of early SARS-CoV-2 variant B.1.1.529 (Omicron) genomic sequences and their epidemiological correlates

**DOI:** 10.1101/2021.12.18.21267908

**Authors:** Ashutosh Kumar, Adil Asghar, Himanshu N. Singh, Muneeb A. Faiq, Sujeet Kumar, Ravi K. Narayan, Gopichand Kumar, Prakhar Dwivedi, Chetan Sahni, Rakesh K. Jha, Maheswari Kulandhasamy, Pranav Prasoon, Kishore Sesham, Kamla Kant, Sada N. Pandey

**Author notes:** **Corresponding author** Department of Anatomy, All India Institute of Medical Sciences (AIIMS), Patna, India, Postal code-801507.

## Abstract

**Background:** A newly emerged SARS-CoV-2 variant B.1.1.529 has worried the health policy makers worldwide due to the presence of a large number of mutations in its genomic sequence, especially in the spike protein region. World Health Organization (WHO) has designated it as a global variant of concern (VOC) and has named as ‘Omicron’. A surge in new COVID-19 cases have been reported from certain geographical locations, primarily in South Africa (SA) following the emergence of Omicron.

**Materials and methods:** We performed an ***in silico*** analysis of the complete genomic sequences of Omicron available on GISAID (until 2021-12-10) to predict the functional impact of the mutations present in this variant on virus-host interactions in terms of viral transmissibility, virulence/lethality, and immune escape. The mutations present at the receptor binding domain (RBD) of the variants were assessed using an open analysis pipeline which integrates a yeast-display platform with deep mutational scanning. Further, we performed a correlation analysis of the relative proportion of the genomic sequences of specific SARS-CoV-2 variants (in the period of 01 Oct-10^th^ Dec, 2021) with the current epidemiological data (new COVID-19 cases and deaths) from SA to understand whether the Omicron has an epidemiological advantage over existing variants.

**Results:** Compared to the current list of global VOCs/VOIs (as per WHO) Omicron bears more sequence variation, specifically in the spike protein and host receptor-binding motif (RBM). Omicron showed the closest nucleotide and protein sequence homology with Alpha variant for the complete sequence as well as for RBM. The mutations were found primarily condensed in spike region (28-48) of the virus. Further, the mutational analysis showed enrichment for the mutations decreasing ACE2-binding affinity and RBD protein expression, in contrast, increasing the propensity of immune escape. An inverse correlation of Omicron with Delta variant was noted (r=-0.99, p< .001, 95% CI: -0.99 to - 0.97) in the sequences reported from SA post-emergence of the new variant, later showing a decrease. There has been a steep rise in the new COVID-19 cases in parallel with increase in the proportion of Omicron since the first case (74-100%), on the contrary, the incidences of new deaths have not been increased (r=-0.04, p>0.05, 95% CI =-0.52 to 0.58).

**Conclusions:** Omicron may have greater immune escape ability than the existing VOCs/VOIs. However, there are no clear indications coming out from the predictive mutational analysis that the Omicron may have higher virulence/lethality than other variants, including Delta. The higher ability for immune escape may be a likely reason for the recent surge in Omicron cases in SA.

**Highlights:**

- Higher immune escape ability than the existing VOCs/VOIs
- No clear indications of increased affinity for ACE2 binding
- Driving a new COVID-19 wave in South Africa
- Outcompeting Delta variant
- Currently, no clear evidence for increased virulence/lethality

## Introduction

A new variant of SARS-CoV-2 (lineage B.1.1.529) has been reported from Botswana, South Africa (SA) and multiple other countries^1^. World Health Organization (WHO) has designated it as a global variant of concern (VOC) and has named as ‘Omicron’^2^. The new variant has been given a pango-lineage as BA.1. Currently very little is known about epidemiological characteristics of this variant. The presence of a large number of mutations in its genomic sequence, especially in the spike protein region including in the host receptor-binding domain (RBD) has raised the speculations that Omicron can prove a serious epidemiological threat and can be a reason for the next COVID-19 waves globally^3^. More recently a sub-lineage (pango-lineage BA.2) has appeared in Omicron with slightly varying set of mutations. We performed an ***in silico*** analysis of the complete genomic sequences, of Omicron available on GISAID, to predict the functional impact of the mutations, present in this variant, on virus-host interactions in terms of viral transmissibility, virulence, and the immune escape capabilities. Further, we made an assesment of the relative proportion of the genomic sequences of the SARS-CoV-2 variants and correlated that with the rise in new COVID-19 cases in a most affected global geographical location by Omicron, as to understand whether the new variant has an epidemiological advantage in terms of transmissibility and virulence/lethality over the existing variants. The findings of this study provides important insights about origin and the epidemiological attributes of this variant paving way for the future research.

## Methods

### Data collection

#### SARS-CoV-2 genomic sequences

The SARS-CoV-2 genomic sequence for Omicron variant and other global variants of concern/interest (VOCs/VOIs) were downloaded from EpiCoVTM database of Global Initiative on Sharing All Influenza Data (GISAID) (https://www.gisaid.org/) using automatic search function feeding information for geographical location, SARS-CoV-2 lineage and sample collection and sequence reporting dates (up to 2021-12-10). The optimum length and coverage of the downloaded sequences (used for variant comparisons) were achieved with selecting ‘complete sequence’ and ‘high coverage’ in the search function.

#### Epidemiological data

The global epidemiological data (daily new cases and deaths) for COVID-19 for the period of 1^st^ Oct-10^th^ Dec, 2021 were accessed from Worldometer (https://www.worldometers.info/coronavirus).

#### Data analysis

The mutational analysis on the genomic sequences was performed and 3D structure of spike with amino acid changes in Omicron was generated using CoVsurver app by GISAID (https://www.gisaid.org/) employing hCoV-19/Wuhan/WIV04/2019 as a reference strain. Further, a comparative mutational analysis of Omicron with existing global VOCs/VOIs (as per WHO)^4^ was generated using ‘Compare lineages’ function at Outbreak.info (https://outbreak.info/) using GISAID as the source of genomic sequence data. The protein sequence translation from the viral genomic sequences were done using Expasy by Swiss Institute of Bioinformatics (https://www.expasy.org/). A comparative assessment of the Omicron nucleotide and protein sequences with existing global VOCs/VOIs were made using NCBI Blast tool (https://blast.ncbi.nlm.nih.gov/Blast.cgi?).

Furthermore, the functional impact of the mutations present at the receptor binding domain (RBD) of the variants were assessed using an open analysis pipeline developed by Starr *et al*, 2020 which integrates a yeast-display platform with deep mutational scanning to determine how all amino acid mutations to measure how all possible RBD amino acid mutations affect ACE2-binding affinity and protein expression (a correlate of protein folding stability) as compared to the wild-type (WT) SARS-CoV-2 strain (https://jbloomlab.github.io/SARS-CoV-2-RBD_DMS/).

The epidemiological correlates of the Omicron variant were assessed based on the comparative analysis of the genomic sequences from GISAID and current epidemiological data (daily new cases and deaths) from one of the most affected region by this variant (last date of collection: 2021-12-10). The number of sequences for each SARS-CoV-2 variants was retrieved using automatic search function feeding information for the lineage and collection dates in EpiCoVTM database of GISAID for the period of 1^st^ Oct, 2021 to 10^th^ Dec, 2021. A 3 day sum of the total number of sequences was noted for each variant and their relative proportions were calculated (in percentage). Data was tabulated, and the distribution of each variant was charted against the COVID-19 epidemiological data (3 day sum of new case and deaths) and statistically analyzed for appreciating the changes in relationship between the variables before and after the emergence of Omicron.

#### Statistics

An expect (E) value ≤ 0 was considered significant for the sequence homology match through NCBI Blast. An *E*-value close to zero or below together with the higher Max score signified a higher sequence homology ranking. (Further details of the statistical methods predicting significance can be consulted at https://www.ncbi.nlm.nih.gov/BLAST/tutorial/Altschul-1.html). For the mutational analysis only the mutations present in atleast 75% of sequenced samples were considered for functional characterisation.

For the analysis of epidemiological data the statistical tests were performed to evaluate inter-group differences between the SARS-CoV-2 Variants with the help of MS Excel 2019 and XLSTAT package. Normality of the data was examined using Shapiro-Wilk test. Pearson’s correlation and Spearman’s rho tests were performed for the normally distributed and skewed data respectively. A correlation matrix was generated and a linear regression analysis was performed between the comparing variables [presented as r values= 0 to 1 at confidence interval (CI) of 95%]. Results were considered statistically significant at p-value ≤ 0.05. Graphs were plotted to visualize the data trends.

#### Ethical statement

An ethical clearance was precluded as *in silico* methods and open access/globally shared databases were used to conduct this study.

## Results

A total of 3604 genomic sequences of Omicron from 54 countries were uploaded on the GISAID until 2021-12-10 (Fig. S1) which were analyzed for the mutational characteristics. The highest number of Omicron genomic sequences (81.5% of the total) were reported from SA where a rise in the COVID-19 cases has been observed post-emergence of the new variant (https://www.worldometers.info/coronavirus/country/south-africa/dated2021-12-10). The mutations found were primarily condensed in spike region (28-48) of the virus, however, frequent non-spike mutations were also noted (20-26). In addition, eight sequences of BA.2 was reported from four countries which included newer mutations in all viral parts including spike protein (Figs. S2-3). In this study, we have focused on analyzing the genomic sequences of currently most prevalent sublineage of Omicron (BA.1).

### Sequence homology of Omicron (BA.1) with WT strains

Compared to the current list of global VOCs/VOIs (as per WHO) Omicron showed more sequence variation, more specifically in the spike protein (n21563-25384: 1-1273aa) including RBM (n22869-23089: 438-508aa), where the variations were most prominent (Table S1). The homology of the Omicron to the reference strain (hCoV-19/Wuhan/WIV04/2019) for the spike protein sequence varied from 96.23 to 97.8% (28-48 mutations) in the analyzed sequences (Fig. 1).

**Figure 1.**
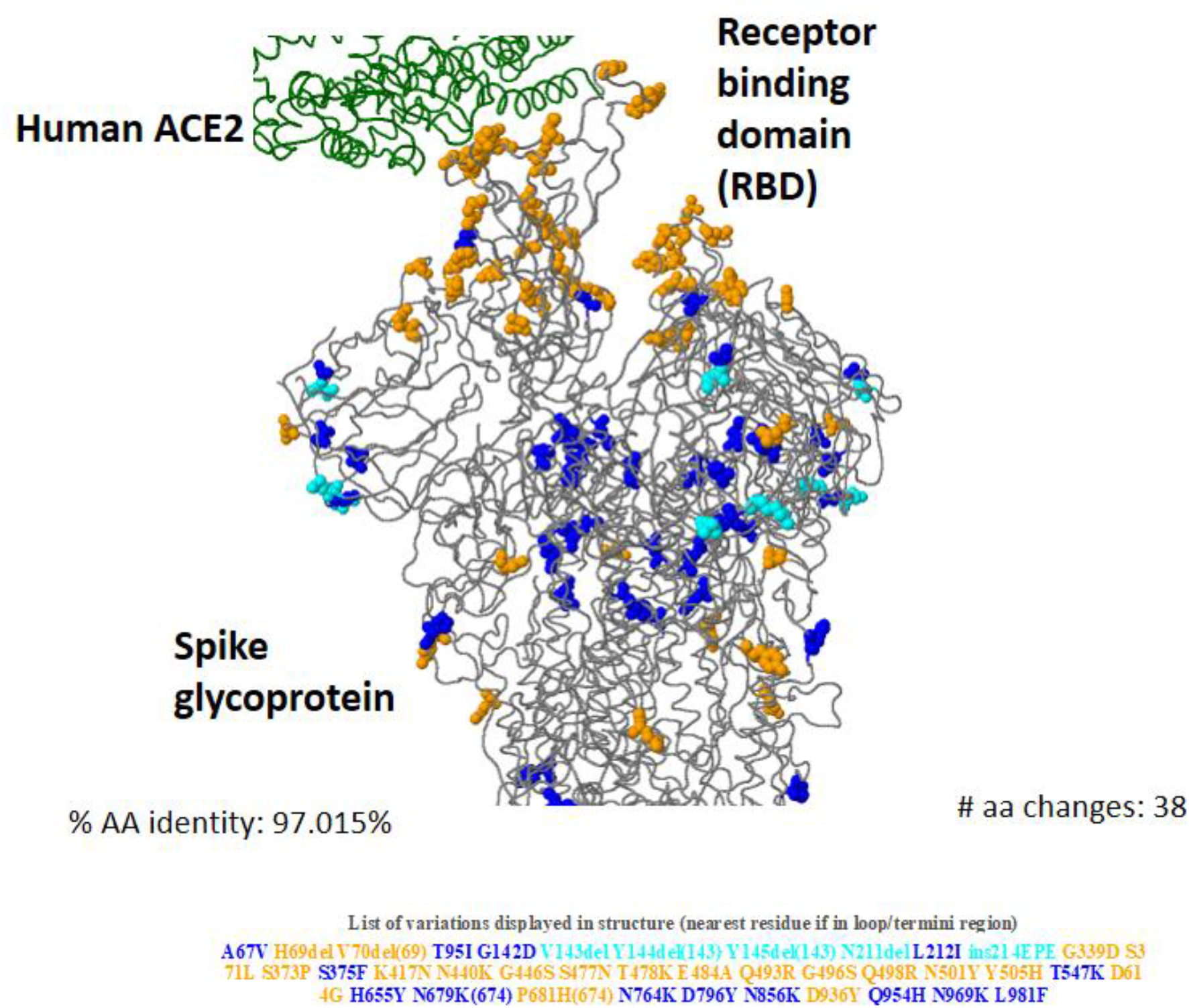
3-D structure of Omicron (BA.1) spike glycoprotein in interaction of human ACE2 showing key amino acid substitutions. (Data source: The figure has been generated from the genomic sequences of Omicron using CoVsurver app from Gisaid.org.)

### Sequence homology of Omicron (BA.1) with existing SARS-CoV-2 VOCs/VOIs

The analysis of genomic and protein sequence homology of Omicron with the reference strain and current global VOCs/VOIs (as per WHO) showed the highest similarity of Omicron with the Alpha variant for the complete sequence as well as RBM, however, the highest similarity for the complete nucleotide and protein sequences for the spike protein were noted with Beta and Delta variants, respectively (Table S1).

### Mutational analysis

The multiple clusters of closely spaced mutations were noted across the sequence, which were most densely placed in the spike protein region, particularly in its S1 subunit, including the host receptor-binding motif (RBM) (Fig. 2, Table 2). Many of the mutations in Omicron are common with the current global VOCs/VOIs (Fig. 3). Table 1 provides a list of the mutations in Omicron (BA.1) (present in at least 75% of sequences) and their functional characteristics based on the existing literature evidence. Based on the available evidence these mutations in pango-lineage BA.1 can be broadly categorized into four major groups: 1. Host receptor binding (10), 2. Host-adaptibility (3), 3. Immune escape (20), and 4. Virus replication (18) (Table 1).

**Figure 2.**
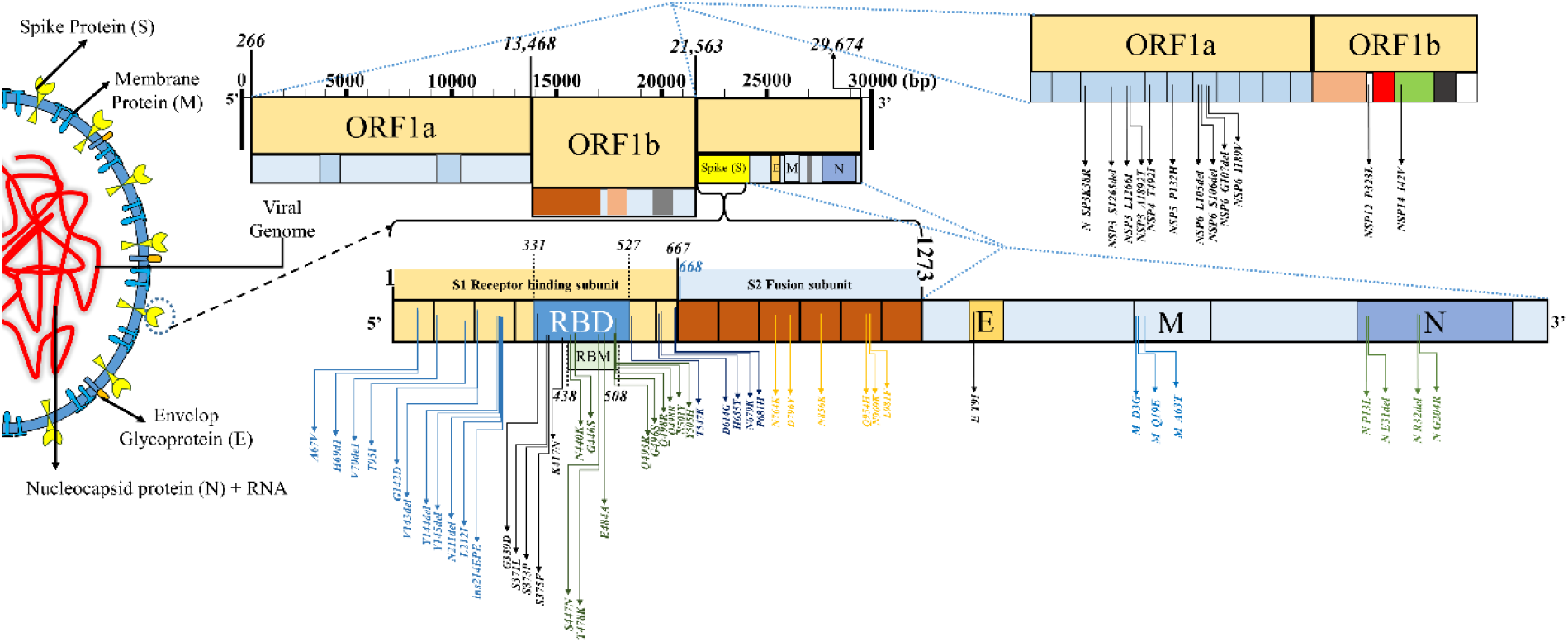
The mutational landscape in SARS-CoV-2 variant B.1.1.529 (Omicron, sublineage: BA.1). Abbreviations: RBD— receptor binding domain, RBM—receptor binding motif, ORF—open reading frame.

**Figure 3.**
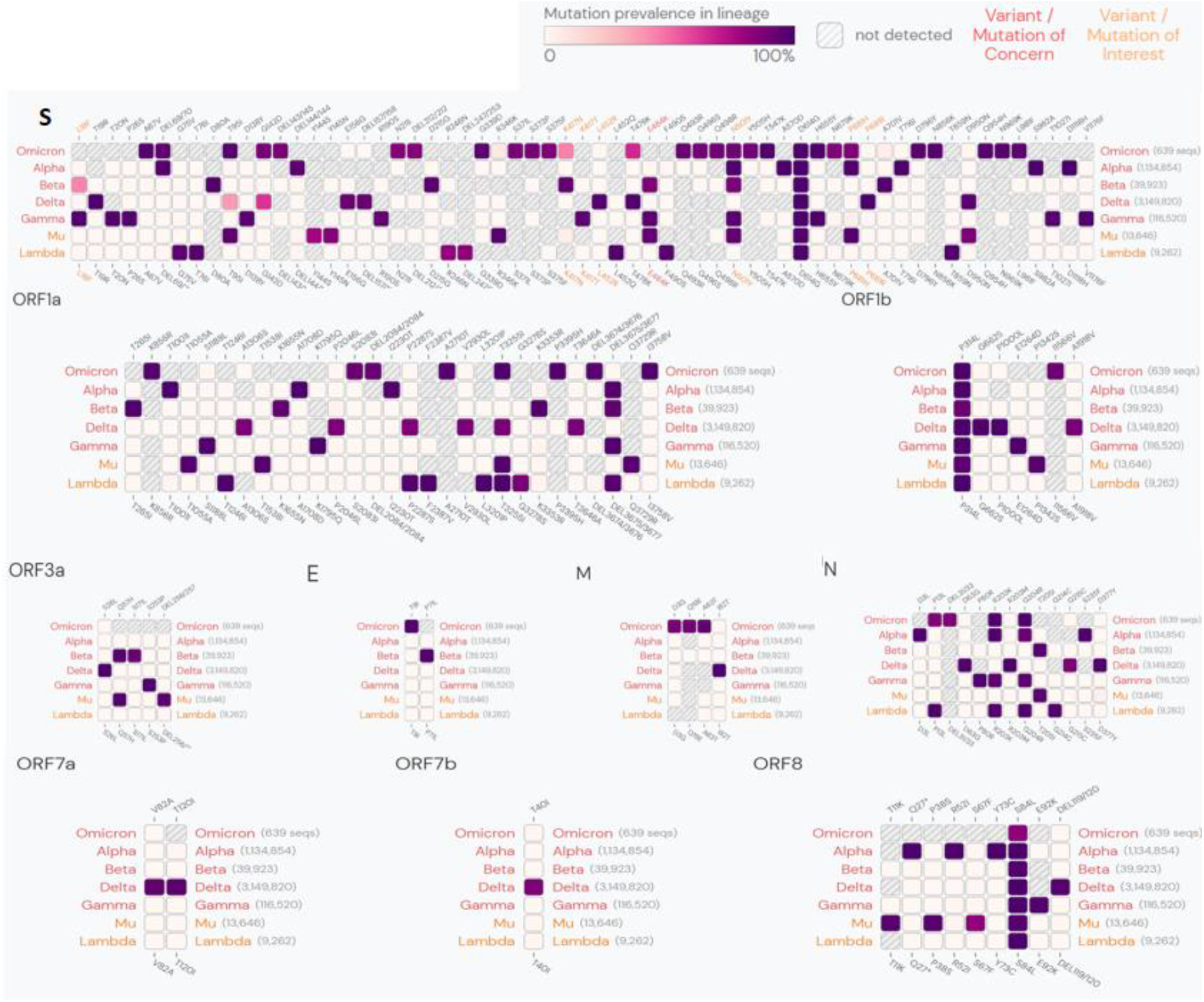
Lineage comparison between Omicron and other global variants of concerns/interest. Showing the mutations with > 75% prevalence in at least one lineage. (Data source: The figure has been generated from outbreak.info, based on the SARS-CoV-2 genomic sequences uploaded in GISAID until 2021-12-6.)

**Table 1.** List of mutations in SARS-CoV-2 variant B.1.1.529 (Omicron, sublineage: BA.1).

| Mutations | Frequency# | Effect on virus-host interactions |  |  |  |  | Remarks | References |
| --- | --- | --- | --- | --- | --- | --- | --- | --- |
|  |  | ACE2 binding/RBD expression | Host adaptability | Immune-escape | Viral Oligomerization Interfaces | Others |  |  |
| A67V | 0.36% | - | - | - | - | - | - | https://www.epicov.org/ |
| H69del | 20.35% | - | - | Antibody Recognition Sites/ Antigenic Drift | - | - | H69del+V70del have 2-fold higher infectivity compared to wildtype.<br>H69del+V70del containing viruses showed reduced neutralisation sensitivity to mAb COVA1-21, targeting an as yet undefined epitope outside the RBD. | https://www.epicov.org/ |
| V70del | 20.37% | - | - | Antibody Recognition Sites/ Antigenic Drift | - | - | H69del+V70del have 2-fold higher infectivity compared to wildtype.<br>H69del+V70del containing viruses showed reduced neutralisation sensitivity to mAb COVA1-21, targeting an as yet undefined epitope outside the RBD. | https://www.epicov.org/ |
| T95I | 21.32% | - | - | - | - | - | - | https://www.epicov.org/ |
| G142D | 33.40% | - | - | - | - | - | - | https://www.epicov.org/ |
| V143del | 0.12% | - | - | Antibody Recognition Sites | - | - | - | https://www.epicov.org/ |
| Y144del | 20.94% | - | - | Antibody Recognition Sites/ Antigenic Drift | - | - | Decreased sensitivity to convalescent sera. | refs<sup>5,6</sup> |
| Y145del | 2.33% | - | - | Antibody Recognition Sites/ Antigenic Drift | - | - | Decreased sensitivity to convalescent sera. | refs<sup>5,6</sup><br>https://www.epicov.org/ |
| N211del | 0.02% | - | - | - | - | - | - | https://www.epicov.org/ |
| L212I | 0.01% | - | - | - | - | - | - | https://www.epicov.org/ |
| ins214EPE | 0.00% | - | - | - | - | - | - | https://www.epicov.org/ |
| G339D | 0.01% | Increased RBD expression | - | Antibody Recognition Sites/ Antigenic Drift | - | - | - | ref<sup>7</sup><br>https://www.epicov.org/ |
| S371L | 0.00% | Increased ACE2 binding | - | Antibody Recognition Sites/ Antigenic Drift | Viral Oligomerization Interfaces | - | - | ref<sup>7</sup><br>https://www.epicov.org/ |
| S373P | 0.01% | Increased RBD expression | - | Antibody Recognition Sites/ Antigenic Drift | Viral Oligomerization Interfaces | - | - | ref<sup>7</sup><br>https://www.epicov.org/ |
| S375F | 0.00% | - | - | Antibody Recognition Sites/ Antigenic Drift | Viral Oligomerization Interfaces | - | - | ref<sup>8</sup><br>https://www.epicov.org/ |
| K417N | 0.83% | Increased RBD expression | - | Antibody Recognition Sites/ Antigenic Drift | Viral Oligomerization Interfaces | - | - | refs<sup>7,9</sup><br>https://www.epicov.org/ |
| N440K | 0.17% | Increased ACE2 binding | - | Antibody Recognition Sites/ Antigenic Drift | - | - | - | ref<sup>7</sup><br>https://www.epicov.org/ |
| G446S | 0.01% | - | - | Antibody Recognition Sites/ Antigenic Drift | - | - | - | ref<sup>9</sup><br>https://www.epicov.org/ |
| S477N | 1.31% | Increased ACE2 binding/ Increased RBD expression | - | Antibody Recognition Sites/ Antigenic Drift | Viral Oligomerization Interfaces | - | S477N was also resistant to neutralization by the human convalescent sera tested in this study, but not to vaccine-elicited sera. | refs<sup>7,10</sup><br>https://www.epicov.org/ |
| T478K | 52.56% | Increased ACE2 binding/ Increased RBD expression | - | - | - | - | Decreased sensitivity to convalescent sera | refs<sup>5,7</sup> |
| E484A | 0.02% | - | - | Antibody Recognition Sites/ Antigenic Drift | - | - | - | ref<sup>8</sup> |
| Q493R | 0.01% | Host change | - | Antibody Recognition Sites/ Antigenic Drift | Viral Oligomerization Interfaces | - | - | refs<sup>11,12</sup> |
| G496S | 0.01% | Increased RBD expression | - | Antibody Recognition Sites/ Antigenic Drift | - | - | - | ref<sup>7</sup> |
| Q498R | 0.00% | - | - | Antibody Recognition Sites/ Antigenic Drift | - | - | - | ref<sup>8</sup><br>https://www.epicov.org/ |
| N501Y | 24.11% | Increased ACE2 binding/Host change | - | Antibody Recognition Sites/ Antigenic Drift | Viral Oligomerization Interfaces | - | N501Y associated with increased transmissibility and increased affinity for human ACE2 receptor. | refs<sup>7,9,13</sup><br>https://www.epicov.org/ |
| Y505H | 0.00% | Increased RBD expression | - | - | Viral Oligomerization Interfaces | - | - | ref<sup>7</sup><br>https://www.epicov.org/ |
| T547K | 0.00% | - | - | - | - | - | - | https://www.epicov.org/ |
| D614G | 98.51% | Increased infectivity | - | - | - | - | D614G mutant increases the infectivity SARS-CoV-2. Lower Ct values were observed in G614 infections indicating higher viral load. | refs<sup>5,14</sup><br>https://www.epicov.org/ |
| H655Y | 2.25% | - | Host adaptation (cats)<br>Spike glycoprotein fusion efficiency | Antibody Recognition Sites/<br>Antigenic Drift | - | - | - | refs<sup>15-17</sup> |
| N679K | 0.09% | - | - | - | - | - | - | ref<sup>18</sup><br>https://www.epicov.org/ |
| P681H | 22.73% | - | - | - | - | - | P681H mutation at the S1/S2 site of the SARS-CoV-2 spike protein may increase its cleavability by furin-like proteases, but that this does not translate into increased virus entry or membrane fusion. | refs<sup>19,20</sup><br>https://www.epicov.org/ |
| N764K | 0.01% | - | - | - | Viral Oligomerization Interfaces | - | - | https://www.epicov.org/ |
| D796Y | 0.08% | - | - | - | Viral Oligomerization Interfaces | - | - | https://www.epicov.org/ |
| N856K | 0.00% | - | - | - | Viral Oligomerization Interfaces | Ligand binding | - | https://www.epicov.org/ |
| Q954H | 0.00% | - | Host adaptation (cell culture) | - | Viral Oligomerization Interfaces | - | - | ref<sup>21</sup><br>https://www.epicov.org/ |
| N969K | 0.00% | - | - | - | Viral Oligomerization Interfaces | - | - | https://www.epicov.org/ |
| L981F | 0.00% | - | - | - | Viral Oligomerization Interfaces | - | - | https://www.epicov.org/ |
| **Non-spike mutations** |  |  |  |  |  |  |  |  |
| E |  |  |  |  |  |  |  |  |
| E T9I | 0.09% |  |  |  | Viral Oligomerization Interfaces |  |  | https://www.epicov.org/ |
| M |  |  |  |  |  |  |  | https://www.epicov.org/ |
| M D3G | 0.08% |  |  |  |  |  |  | https://www.epicov.org/ |
| M Q19E | 0.00% |  |  |  |  |  |  | https://www.epicov.org/ |
| M A63T | 0.01% |  |  |  |  |  |  | https://www.epicov.org/ |
| N |  |  |  |  |  |  |  |  |
| N P13L | 0.63% |  |  | Antigenic drift |  |  | P13L variant in the B*27:05-restricted CD8+ nucleocapsid epitope showed complete loss of responsiveness to the T-cell lines evaluated | ref<sup>22</sup><br>https://www.epicov.org/ |
| N E31del | 0.00% |  |  |  |  |  |  | https://www.epicov.org/ |
| N R32del | 0.00% |  |  |  |  |  |  | https://www.epicov.org/ |
| N G204R | 26.20% |  |  |  |  |  |  | https://www.epicov.org/ |
| NSP |  |  |  |  |  |  |  | https://www.epicov.org/ |
| NSP3 K38R | 0.01% |  |  |  |  |  |  | https://www.epicov.org/ |
| NSP3 S1265del | 0.02% |  |  |  |  |  |  | https://www.epicov.org/ |
| NSP3 L1266I | 0.02% |  |  |  |  |  |  | https://www.epicov.org/ |
| NSP3 A1892T | 0.00% |  |  |  |  |  |  | https://www.epicov.org/ |
| NSP4 T492I | 47.76% |  |  |  | Viral Oligomerization Interfaces |  |  | https://www.epicov.org/ |
| NSP5 P132H | 0.01% |  |  |  |  |  |  | https://www.epicov.org/ |
| NSP6 L105del | 0.02% |  |  |  |  |  |  | https://www.epicov.org/ |
| NSP6 S106del | 24.74% |  |  |  |  |  |  | https://www.epicov.org/ |
| NSP6 G107del | 24.74% |  |  |  |  |  |  | https://www.epicov.org/ |
| NSP6 I189V | 0.03% |  |  |  |  |  |  | https://www.epicov.org/ |
| NSP12 P323L | 96.69% |  |  |  | Viral Oligomerization Interfaces |  |  | https://www.epicov.org/ |
| NSP14 I42V | 0.00% |  |  |  | Viral Oligomerization Interfaces |  |  | https://www.epicov.org/ |

\*Based on the genomic sequences of Omicron uploaded on the GISAID.ORG (last date of collection 2021-12-10).
#Among all SARS-CoV-2 genomic sequences uploaded on GISAID.ORG.

**Table 2.** The predicted impact of receptor binding motif (RBM) variations in SARS-CoV-2 variant B.1.1.529.

| ACE2 binding site mutations | ACE2 binding ( $\Delta\log_{10}$ KD,app *) | Protein expression ($\Delta\log$ mean MFI#) | ACE2: SARS-CoV-2 contact | RSA bound | SARS-CoV-1-aa | RaTG13-aa | GD Pangoline-aa |
| --- | --- | --- | --- | --- | --- | --- | --- |
| G339D | 0.06 | 0.30 | false | 0.47 | G | G | G |
| S371L | -0.14 | -0.61 | false | 0.46 | S | S | S |
| S373P | -0.08 | -0.22 | false | 0.48 | F | S | S |
| S375F | -0.55 | -1.81 | false | 0.48 | S | S | S |
| K417N | -0.45 | 0.10 | true | 0.19 | V | K | R |
| N440K | 0.07 | -0.12 | false | 0.68 | N | H | N |
| G446S | -0.20 | -0.40 | true | 0.55 | T | G | G |
| S477N | 0.06 | 0.06 | false | 0.76 | G | S | S |
| T478K | 0.02 | 0.02 | false | 0.48 | K | K | T |
| E484A | -0.07 | -0.23 | false | 0.5 | P | T | E |
| Q493R | -0.09 | -0.06 | true | 0.1 | N | Y | Q |
| G496S | -0.63 | 0.12 | true | 0.04 | G | G | G |
| Q498R | -0.06 | 0.10 | true | 0 | Y | Y | H |
| N501Y | 0.24 | -0.14 | true | 0.03 | T | D | N |
| Y505H | -0.71 | 0.16 | true | 0.12 | Y | H | Y |

(Omicron, sublineage: BA.1) on interactions with host<sup>§</sup>.
<sup>§</sup> Based on study by Starr et al., (2020)<sup>7</sup> \*A positive $\Delta\log_{10}$ KD,app value relative to the unmutated SARS-CoV-2 RBD ( $3.9 \times 10^{-11}$ M ) indicates stronger binding. # $\Delta\log$ MFI relative to the unmutated SARS-CoV-2 RBD-positive values indicating increased expression.
**Abbreviations:** KD,app— apparent dissociation constants, MFI— mean fluorescence intensity, RSA-relative solvent accessibility.

The analysis of the mutations present at the RBD using deep mutational scanning pipeline by Starr *et al*, 2020 reflected prominent ACE2-binding affinity and protein expression changes (Table 2). Notably, mutations decresing the ACE2-binding affinity and protein expression were found significantly greater in number.

### Epidemiological correlates

A total (N) of 4224 SARS-CoV-2 genomic sequences (NDelta = 999, NOmicron = 2937, and NOthers = 288) were uploaded on GISAID from SA in the period of study. For the complete duration of the study Delta correlated negatively with the new COVID-19 cases (r=-0.567, p=0.004, 95% CI=-0.79 to - 0.21), however positively with the new deaths (r=0.38, p=0.065, 95% CI= -0.025 to 0.68). The differential analysis of the SARS-CoV-2 genomic sequences from SA prior and after emrgence of the first case of Omicron (dated 2021-11-05, EPI_ISL_7456440) reflected sharp change in the dominance of the variant from Delta to Omicron (Fig. 4). An inverse correlation of Omicron with Delta variant was noted (r=-0.99, p< .001, 95% CI: -0.99 to -0.97) in the period of study. There has been a steep rise in the new COVID-19 cases in parrallel with the increase in proportion of Omicron since the first case of Omicron (74-100% of total genomic sequences after 15-17^th^ Nov, 2021), however, no parrallel increase has been observed in the death cases, which otherwise showed a reverse trend (r=-0.04, p>0.05, 95% CI =-0.52 to 0.58) (Fig. 4).

**Figure 4.**
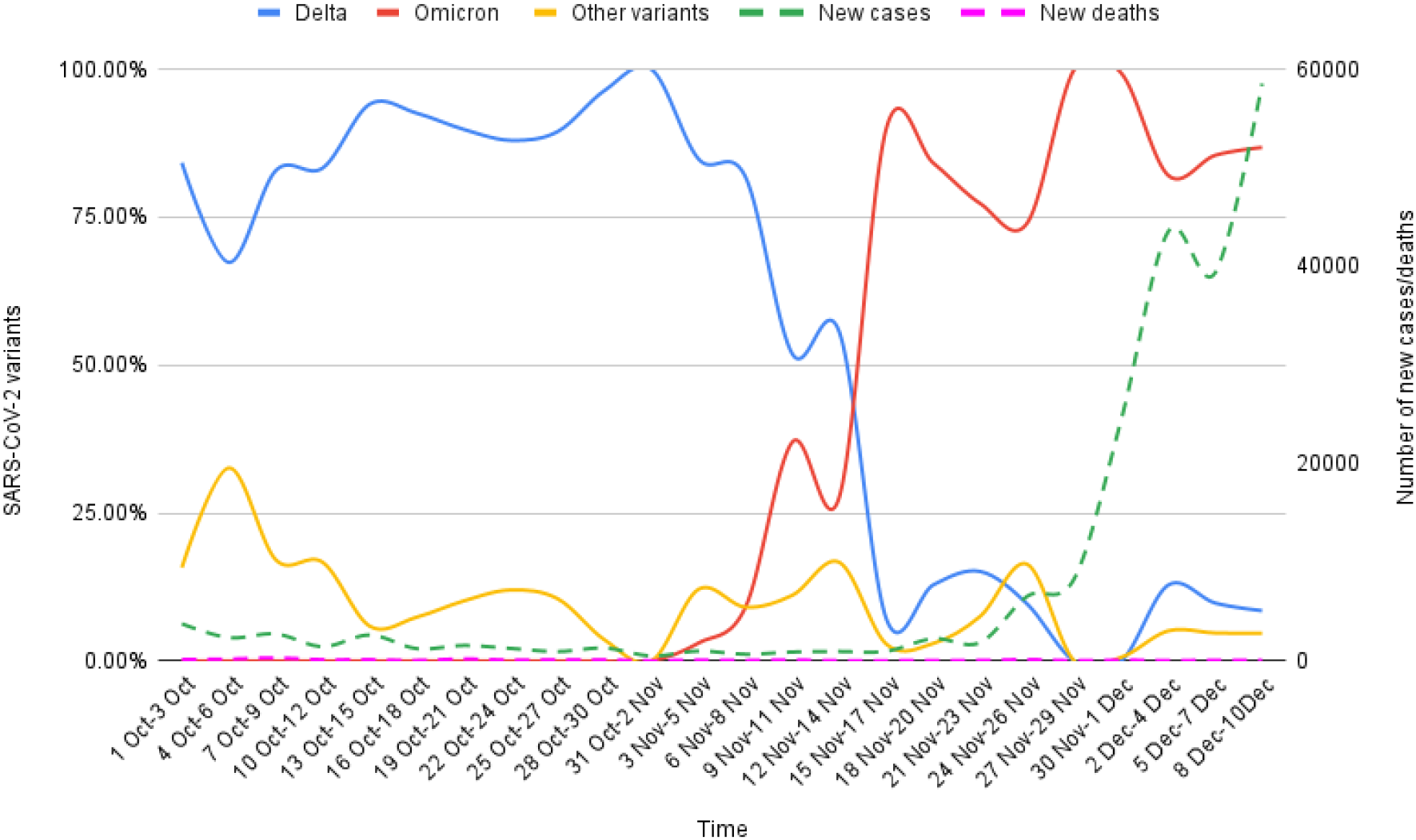
Epidemiological correlates of Omicron and Delta variants genomic sequences reported on GISAID from South Africa for the period of 1^st^ Oct-10^th^ Dec, 2021. The proportion of Delta and Omicron variants in the total SARS-CoV-2 genomic sequences were correlated with the new COVID-19 cases and deaths in the period of study (3-day sum of each variable). A sharp change in the dominance from Delta to Omicron is observable since the first case (5^th^ Nov, 2021). The rise of Omicron cases has parrelled with the rise in the new COVID-19 cases. In comparison, the Delta variant is showing a fall in the same time period. Notably, there has been no increase in the number of deaths post emergence of Omicron. (Data source: gisaid.org and worldometer.info.)

## Discussion

Our analysis of the SARS-CoV-2 genomic sequences coupled with epidemiological data from SA unravels multiple observations regarding host-virus interactions which may help predicting the epidemiological potentials of the Omicron.

### Sequence variation with WT strains and existing SARS-CoV-2 VOCs/VOIs

Our analyses showed that Omicron has significantly greater variantions than current VOCs and VOIs in comparisons with the wild type strains (Wuhan strain and B.1). The sequence variation was greatest in the spike protein region, more particularly in the receptor-binding motif (RBM). Among the existing VOCs and VOIs, the highest homology of the complete sequence and RBM (nucleotide as well as protein) of Omicron was observed with Alpha variant (Table S1). Interestingly, similar to Alpha variant S-gene target failure (SGTF) in PCR based detection is proving a sensitive method for the detection of Omicron in the clinical samples^23^.

As Omicron bears key mutations from multiple existing VOCs/VOIs, with approximate sequence homology variation, rather than a direct descendency, the multiple recombination events between the variants inside the hosts can be a more plausible explanation for its origin. Notably, compared with the WT strains, Omicron has significantly higher sequence variation than existing VOCs/VOIs, more specifically in spike protein region (Table S1). Further, in the spike protein region, the highest sequence variations is noted in RBM (Table S1).

It will be pertinent to explore the evolutionary mechanisms involved in accumulation of such a large number of mutations in Omicron. The speculations have also been raised that the long term persistence of SARS-CoV-2 infection in an immunocompromised subject can be a probable mechanism behind the Omicron origin. Existing literature^24–27^ supports strong plausibility of this mechanism. Avanzato *et al*^24^ and Choi *et al*^26^ reported case studies of persistence of infection and accumulation of novel mutations in SARS-CoV-2 spike gene and RBD in a chronically ill and immunocompromised COVID-19 patients. Another such case has been recently reported by Karim *et al*^25^. Authors documented long persistence of SARS-CoV-2 infection (more than six month) in a patient with advanced HIV and antiretroviral treatment failure. Through whole genome sequencing for SARS-CoV-2, performed at multiple time-points from the patient samples, authors demonstrated early emergence of the E484K substitution followed by N501Y, K417T, and multiple others mutation (including some novel mutations) in the spike gene and RBD. Increase in the genomic diversity reflecting intra-host evolution of SARS-CoV-2 during prolonged infection was also noted in a recent cohort study by Voloch *et al*^27^.

### Predicted effect on virus-host interactions

The new variant has accumulated multiple closely spaced mutations at receptor-binding motif (RBM) with ACE2, specifically a cluster of mutations have appeared at 477-501aa (Fig. 2). Notably, it has many of the mutations common with the earlier variants of concern (Fig. 3), many of which have been shown to enhance RBD:ACE2 binding in comparison to the WT strain^28^, as has also been reflected in our analysis (Table 1-2). The selective mutations which are either present at or in the vicinity of the RBM (N440K, S477N, T478K, and N501Y) in majority of the Omicron sequences are believed to stabilize binding with ACE2 (Table 2). D614G, a key mutation characteristically present in all B.1 descendents^28^, is known to stabilize the trimeric structure and create a more open conformation of the RBD allowing a stronger binding with ACE2^28^.

In paradox, our analysis shows that the majority of the novel or rare spike mutations (<0.2% prevalence in the total sequenced samples, Table 1) in Omicron may have deleterious effect on the host interactions owing to their presence at the constrained RBD regions in terms of ACE2 binding (10/15) and/or RBD expression (8/15) (Table 2). Notably, most of the spike mutations which predicted a favorable effect on ACE2 binding or RBD expression or both are present in current VOCs, primarily Delta (T478K), and Alpha (N501Y), Beta (K417N) variants. Further, a set of mutations in Omicron which are present inside (P681H) or in vicinity (D614G, H655Y) of the furin cleavage site (FCS) of SARS-CoV-2 spike protein — a small stretch of peptide (PRRAR) inserted at the intersection of spike segments S1 and S2 (681–685 aa residues)— can enhance proteolytic cleavage of spike protein by a host protease-furin, which is said to improve its fusion to the host-cell membrane^29^. P681H is characteristically present in multiple VOCs/VOIs such as B.1.1.7, P.1, Q.1 and B.1.621 lineage variants^30^. A mutation at the same location P681R has been present in Delta variant and its emerging sub-lineages^31^. The characterization of the individual mutations present on RBM gives a speculation that Omicron may not have more efficient interactions with the host than existing VOCs/VOIs, more specifically Delta. A further assessment of the allosteric influence and dynamic interactions of the mutations present at RBD and other regions of spike protein, and examining in *in situ*/*vivo* studies will be necessary to understand their exact impact on host-receptor binding and its clinical correlates. The clinical data on severity of the illness in Omicron is currently very limited, although the evaluation of early cases has indicated a milder illness compared to the existing VOCs^32^.

### Viral replication

Many of the mutations, especially in the non-spike regions, are linked with viral oligomerization and synthesis and packaging of the ribonucleic acid core (Table 1) and it is likely that they may have a role in replication of the virus inside the host cells^33^. Particular to notice has been NSP12 P323L, a mutation present in *RNA-dependent RNA polymerase (RdRp)* coding region (Fig. 1, Table 1). Of note this has been a frequently observed mutation in the earlier variants (96.69%) (Table 1). However, whether these mutations will have a positive or a negative impact on the viral replication is little understood yet. Interestingly, the results of a preliminary study^34^ which employed *ex-vivo* cultures of Omicron isolated from the respiratory tract of the infected patients, and compared that with WT and Delta variant strains. The authors observed that after 24 hours of incubation Omicron replicated 70 times faster than WT and Delta variant strains in the human bronchus. In contrast, it replicated less efficiently (>10 times lower) in the human lung tissue than WT strain, and also lower than Delta variant.

### Immune escape

Majority of the spike mutations (18/32) in Omicron have occurred at the known antibody recognition sites (Table 1). Existing studies have established role of these mutations in immune escape against convalescent sera, vaccine acquired antibodies, and therapeutically used monoclonal antibodies (Table 1). The emerging preliminary evidence from *in situ* studies indicate potential immune escape by Omicron against convalescent sera, vaccine acquired antibodies, and therapeutically used monoclonal antibodies ^23,35,36^. Interestingly, Omicron contains K417N and E484A, which are present in multiple existing variants, and are believed to contribute to immune escape^28^. Of note, K417 locus is a known epitope for CB26—a therapeutically used monoclonal antibody in COVID-19^28^. A larger number of mutations in Omicron spike protein, more specifically in RBD may be an evolutionary gain in this variant providing it higher immune escape. A support for this notion comes from a recent study by Nabel *et al*^37^ who demonstrated that SARS-CoV-2 pseudotypes containing up to seven mutations, as opposed to the one to three found in earlier variants of concern, are more resistant to neutralization by therapeutic antibodies and serum from vaccine recipients.

A non-spike mutation (N) P13L present in Omicron (Table 1) is shown to cause complete loss of recognition by epitope-specific (B*27:05-restricted CD8+ nucleocapsid epitope QRNAPRITF9-17) T cells in a recent cell-line based *in situ* study^22^, however, no such evidence in human samples are currently available. In a recent preliminary study Redd *et al* examined peripheral blood mononuclear cell (PBMC) samples from PCR-confirmed, recovered/ convalescent COVID-19 cases (n=30) for their anti-SARS-CoV-2 CD8+ T-cell responses with Omicron. Authors noted that only one low-prevalence (found in 7%) epitope (GVYFASTEK, restricted to HLA*A03:01 and HLA*A11:01) from the spike protein (T95I) region was mutated in Omicron^38^. Presence of these mutations raises concerns about escaping T cell immunity by Omicron^39^ and hence are required to be explored in detail.

Currently *in situ/vivo* studies are scarce which have examined Omicron response against current COVID-19 vaccines in use, although, emerging preliminary evidence is supporting the prediction of immune escape for this variant^23,35,36,40^. Cele *et al*^23^ tested the ability of plasma from 14 BNT162b2 vaccinated study participants to neutralize Omicron versus WT D614G virus in a live virus neutralization assay.The authors observed that Omicron showed a 41-fold decline in FRNT50 Geometric mean titer (GMT) compared to WT D614G virus in the subjects without previous infection (6/14). Interestingly, those with previous infection showed relatively higher neutralization titers with Omicron (6/14), which indicated that the previous infection, followed by vaccination or booster may increase the levels of neutralization and protection from severe disease in cases of Omicron infection.

### Epidemiological correlates: Omicron vs. Delta variants

The analysis of the SARS-CoV-2 genomic sequences from SA indicates the Omicron has gained an advantage in terms of transmissibility over Delta variant (Fig. 4). A third COVID-19 wave driven by Delta variant has recently occurred in SA^41^ hence current epidemiological characteristics of Delta and newly emerged variant in the local population should be analyzed in this backdrop. Our analyses showed that before the arrival of Omicron, Delta variant was dominant locally, on the contrary, at present majority of the new sequences are from Omicron (Omicron vs. Delta: r=-0.99, p< .001, 95% CI: -0.99 to -0.97) (Fig. 4). The steep rise in the new COVID-19 cases in SA are seem to be driven by Omicron, in contrast, Delta variant linked cases are seeing a decline (Fig. 4). The rapid rise in the new COVID-19 cases linked with the emergence of a new SARS-CoV-2 variant is strongly indicating the commencement of a new COVID-19 wave in SA^42^.

Further, the death, which is considered a strong indicator of the virulence/lethality, is showing a negative correlation (r=-0.04, p>0.05, 95% CI =-0.52 to 0.58) (Fig. 4) with rise in Omicron, however, it correlates positively with Delta variant in the period post-emergence (r=0.38, p=0.065, 95% CI= - 0.025 to 0.68) in the complete duration of the study. The pattern indicates that current deaths are primarily linked with Delta rather than Omicron.

At present, the studies are scarce which have examined transmissibility advantage for Omicron. A ∼2.4 (2.0-2.7) times higher transmissibility with Omicron compared to Delta variant in Gauteng, SA was indicated in a recent conference presentation by South African COVID-19 Modelling Consortium^43^. Another preliminary estimate from UK has indicated that with Omicron risk of spreading infection to the members in a household is three times higher compared to Delta variant^44^. Based on the current epidemiological patterns observed in SA, an epidemiological advantage to Omicron in comparison to Delta can be inferred in terms of transmissibility, however, our analysis has not indicated for increased lethality with Omicron compared to Delta and other variants currently circulating in SA population. However, it will be too early to assess the virulence/lethality with Omicron and a study of epidemiological patterns over a longer time period will be mandatory to develop a conclusive view on this.

It is noteworthy that the presence of an immunological barrier in the population imparted by recent COVID-19 wave mediated by Delta variant may be a likely reason for fall in new cases by this variant. A continuous fall in Delta cases is also noticeable in the period prior to emergence of Omicron (Fig. 4), which is further substantiating this notion. The data records show that a significant proportion of the local population in SA has been fully vaccinated by now (25.2%)^45^. Notably, the high number of immune escape mutations in Delta has contributed to lowered efficacy of the vaccines, immunity from the natural infections, and therapeutically used antibodies^28^. As Omicron contains much higher number of immune escape related mutations, including many shared with Delta (Fig. 3), it is likely that Omicron may have added potential for the vaccine breakthorugh infections and reinfections. Any supportive evidence in literature which examined immune escape capability of Omicron in epidemiological data is currently scarce, however, similar speculations have been presented by few other authors and the global health regulatory bodies^2,32,46^. Interestingly, a higher risk of reinfections with Omicron has been indicated in a recent preprint article by Pulliam *et al*^47^. The authors of this study have performed a retrospective analysis of routine epidemiological surveillance data to examine whether SARS-CoV-2 reinfection risk has changed through time in SA, in the context of the emergence of the consecutive variants: Beta, Delta, and Omicron. The authors noted that as compared to the first wave driven by WT strains subsequent waves by Beta and Delta variant had lower estimated hazard ratio for reinfection versus primary infection [relative hazard ratio for wave 2 vs. wave 1: 0.75 (CI95: 0.59-0.97); for wave 3 vs. wave 1:0.71 (CI95: 0.56-0.92)] in comparison to Omicron [Omicron surge vs. wave 1: 2.39 (95%CI: 1.88-3.11)].

### Conclusion

***In silico*** analysis of the viral genomic sequences suggest that the Omicron may have greater immune escape ability than the existing VOCs/VOIs including Delta. There are no clear indications that Omicron may have higher virulence/lethality than other, so far reported variants. Therefore, the higher ability for immune escape can be a likely reason for the recent surge in Omicron cases in SA.

## Limitations

The limitations of this study needs to be considered while interpreting its clinical and epidemiological significance. Firstly, as of now very limited number of genomic sequences and epidemiological data from the geographical regions affected with Omicron is available for the analysis, which may have an impact on the results. It is likely that the relative frequency of certain lineage characterising mutations in the Omicron variant may vary in future, as more number of sequences are accumulated over time. The epidemiological correlates for the new variant are only indicative of current preliminary trends and requires to be followed for a longer period, more specifically the deaths.

## Data Availability

The data generated in this study has been included with the submission.

## Data sharing

Primary data used for this study are publicly available on: SARS-CoV-2 genomic sequence—GISAID database: https://www.gisaid.org/; Epidemiological data—Worldometer: https://www.worldometers.info/coronavirus/coronavirus/country/india). The categorized data for the study period can be availed from the corresponding author on reasonable request.

## Acknowledgements

The study has used SARS-CoV-2 genomic sequence and epidemiological data from GISAID (https://www.gisaid.org/), outbreak.info (http://outbreak.info/), and Worldometer (https://www.worldometers.info/coronavirus/coronavirus/country/india), respectively.

## Author (s) contributions

AK, GK, and PD collected samples and analyzed data. AK wrote first draft. AA and HS performed statistical analysis. MF, SK, RKN, RKJ, CS, MK, PP, KS, KK, and SNP reviewed and edited the paper. All authors consented for submitting final draft.

## Financial support

None

## Conflict of Interest

None

## Notes

### Competing Interest Statement

The authors have declared no competing interest.

### Funding Statement

This study did not receive any funding

### Author Declarations

1. Global Initiative on Sharing All Influenza Data (GISAID) (https://www.gisaid.org/) 2. Outbreak.info (https://outbreak.info/) 3. Worldometer-South Africa (https://www.worldometers.info/coronavirus/country/south-africa/)

