## Supplementary material for "An *in silico* analysis of early SARS-CoV-2 variant B.1.1.529 (Omicron) genomic sequences and their epidemiological correlates": Figs. S1-3, and Table S1

**Kumar et al., 2021**

Supplementary data: Figs.-S1-3, Table S1.

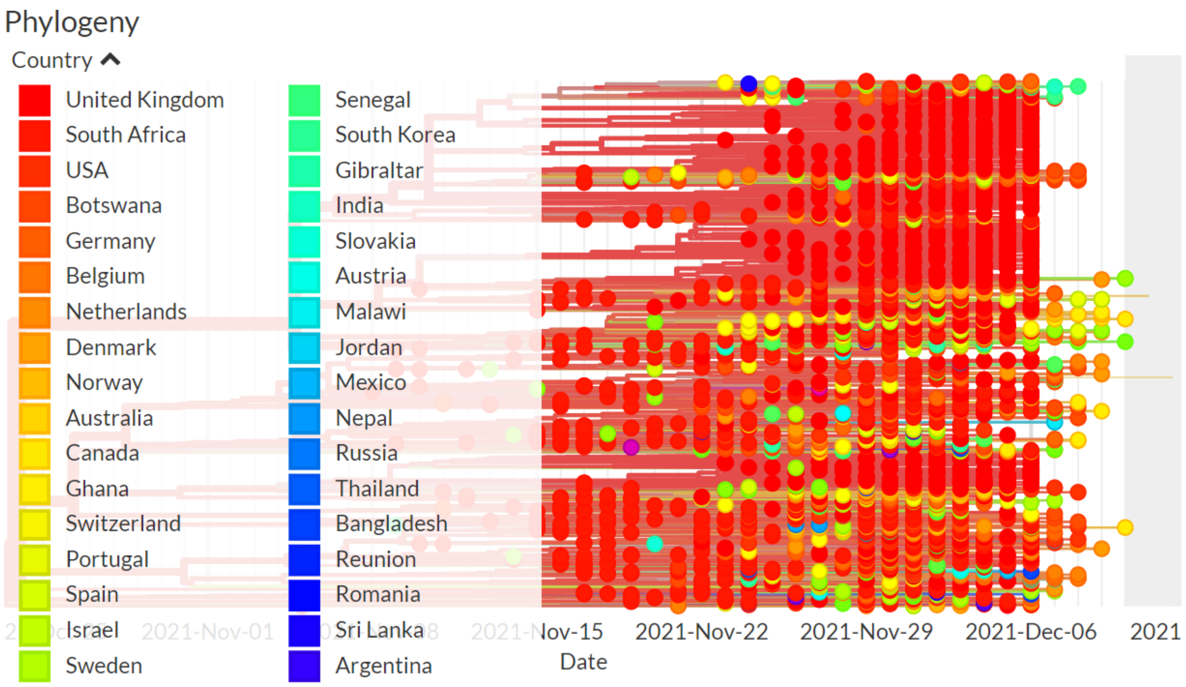


Figure S1 **Global spread of SARS-CoV-2 Omicron variant ( up to 10th Dec, 2021).** (Data source: The figure has been generated from the metadata provided with genomic sequences of Omicron using CoVsurver app from Gisaid.org.)

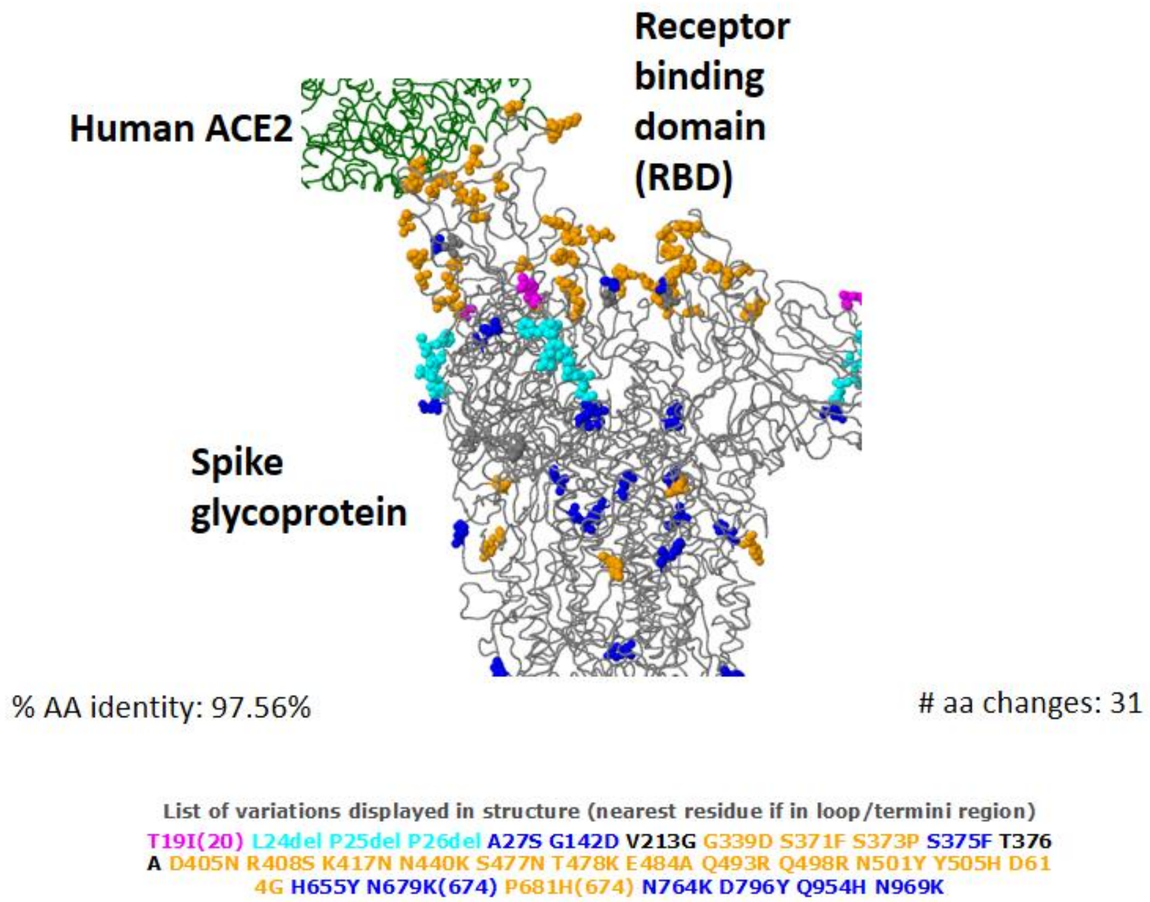


Figure S2 **3-D structure of Omicron (BA.2) spike glycoprotein in interaction of human ACE2 showing key amino acid substitutions.** (Data source: The figure has been generated from the genomic sequences of Omicron using CoVsurver app from Gisaid.org.)

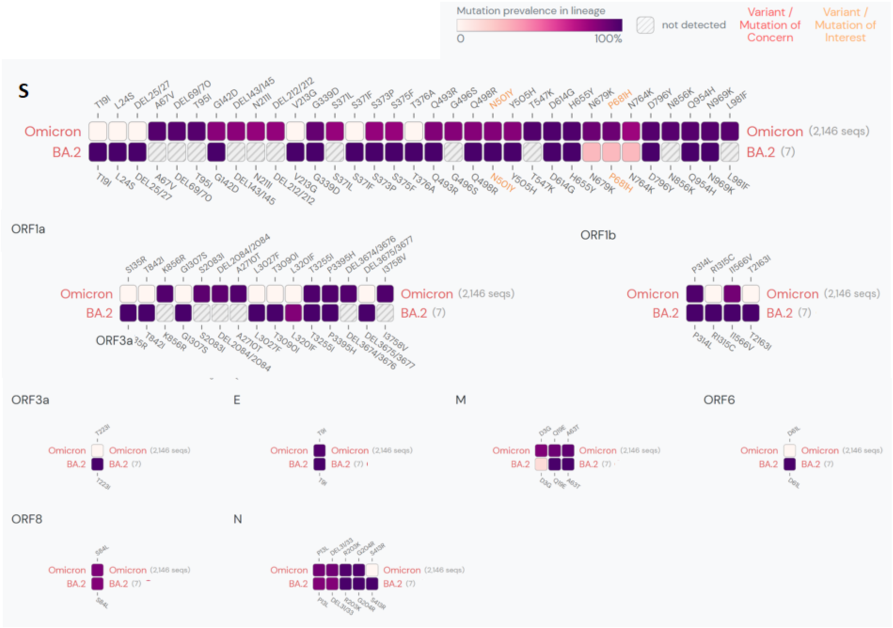


Figure S3 **Lineage comparison between Omicron (BA.1) and its sub-lineage BA.2. Showing the mutations with > 75% prevalence in at least one lineage. Figure has been generated from outbreak.info based on the SARS-CoV-2 genomic sequences uploaded in GISAID until 2021-12-10.**

Table S1 **Nucleotide and protein sequence homology of Omicron (BA.1)* with wild type SARS-CoV-2 and current global variants of concern/interest strains (WHO).**

### S1. 1 Nucleotide sequence alignment

#### S1.1.1 Complete nucleotide sequence

| **Ranking** | **SARS-CoV-2 strain** | **Max score** | **Total score** | **Query cover** | **E value** | **Percentage Identity** | **Accession length** | **GISAID/NCBI Accession number** |
|---|---|---|---|---|---|---|---|---|
| 1. | Alpha | 25401 | 53371 | 97% | 0.0 | 99.75 | 29845 | EPI_ISL_6865096 |
| 2. | Wild type (WT) | 25351 | 53419 | 97% | 0.0 | 99.67 | 29903 | NC_045512.2 |
| 3. | WT with D614G mutation (B.1) | 25346 | 53411 | 97% | 0.0 | 99.66 | 29996 | EPI_ISL_6832722 |
| 4. | Beta | 25298 | 53144 | 97% | 0.0 | 99.62 | 29720 | EPI_ISL_6838995 |
| 5. | Mu | 25292 | 53258 | 97% | 0.0 | 99.59 | 29823 | EPI_ISL_6811664 |
| 6. | Lambda | 25207 | 53221 | 97% | 0.0 | 99.52 | 29856 | EPI_ISL_6694892 |
| 7. | Gamma | 19038 | 51069 | 93% | 0.0 | 99.89 | 29859 | EPI_ISL_6864534 |
| 8. | Delta | 18979 | 52598 | 96% | 0.0 | 99.78 | 29836 | EPI_ISL_6864866 |

#### S1.1.2 Spike nucleotide sequence

| **Ranking** | **SARS-CoV-2 strain** | **Max score** | **Total score** | **Query cover** | **E value** | **Percentage Identity** | **Accession length** | **GISAID/NCBI Accession number** |
|---|---|---|---|---|---|---|---|---|
| 1. | Beta | 2876 | 6368 | 94% | 0.0 | 99.62 | 29720 | EPI_ISL_6838995 |
| 2. | WT with D614G mutation (B.1) | 2876 | 6442 | 94% | 0.0 | 99.62 | 29996 | EPI_ISL_6832722 |
| 3. | Wild type (WT) | 2876 | 6437 | 94% | 0.0 | 99.62 | 29903 | NC_045512.2 |
| 4. | Mu | 2870 | 6429 | 94% | 0.0 | 99.56 | 29823 | EPI_ISL_6811664 |
| 5. | Lambda | 2870 | 6274 | 94% | 0.0 | 99.56 | 29856 | EPI_ISL_6694892 |
| 6. | Alpha | 2865 | 6462 | 94% | 0.0 | 99.49 | 29845 | EPI_ISL_6865096 |
| 7. | Delta | 2865 | 6392 | 94% | 0.0 | 99.49 | 29836 | EPI_ISL_6864866 |
| 8. | Gamma | 2854 | 6398 | 94% | 0.0 | 99.39 | 29859 | EPI_ISL_6864534 |

#### S1.1.3 Receptor binding motif (RBM) nucleotide sequence

| **Ranking** | **SARS-CoV-2 strain** | **Max score** | **Total score** | **Query cover** | **E value** | **Percentage Identity** | **Accession length** | **GISAID/NCBI Accession number** |
|---|---|---|---|---|---|---|---|---|
| 1. | Alpha | 313 | 313 | 85% | 3e-87 | 95.32 | 29845 | EPI_ISL_6865096 |
| 2. | Delta | 313 | 313 | 85% | 3e-87 | 95.32 | 29836 | EPI_ISL_6864866 |
| 3. | Mu | 307 | 307 | 85% | 1e-87 | 95.79 | 29823 | EPI_ISL_6811664 |
| 4. | Gamma | 307 | 307 | 85% | 1e-87 | 95.79 | 29859 | EPI_ISL_6864534 |
| 5. | Beta | 307 | 307 | 85% | 1e-87 | 95.79 | 29720 | EPI_ISL_6838995 |
| 6. | WT with D614G mutation (B.1) | 307 | 307 | 85% | 1e-87 | 95.79 | 29996 | EPI_ISL_6832722 |
| 7. | Wild type (WT) | 307 | 307 | 85% | 1e-87 | 95.79 | 29903 | NC_045512.2 |
| 8. | Lambda | 302 | 302 | 85% | 5e-87 | 95.26 | 29856 | EPI_ISL_6694892 |

### S1. 2 Protein sequence alignment

#### S1.2.1 Complete protein sequence

| Ranking | SARS-CoV-2 strain | Max score | Total score | Query cover | E value | Percentage Identity | Accession length | GISAID/NCBI Accession number |
|---|---|---|---|---|---|---|---|---|
| 1. | Alpha | 56353 | 1.139e+05 | 100% | 0.0 | 99.05 | 55890 | EPI_ISL_6865096 |
| 2. | Beta | 25298 | 1.138e+05 | 99% | 0.0 | 99.21 | 55693 | EPI_ISL_6838995 |
| 3. | Wild type (WT) | 25351 | 1.131e+05 | 100% | 0.0 | 98.22 | 56040 | NC_045512.2 |
| 4. | Lambda | 25207 | 1.137e+05 | 100% | 0.0 | 98.82 | 55954 | EPI_ISL_6694892 |
| 5. | WT with D614G mutation (B.1) | 25346 | 1.141e+05 | 100% | 0.0 | 99.47 | 56235 | EPI_ISL_6832722 |
| 6. | Mu | 25292 | 1.139e+05 | 99% | 0.0 | 99.40 | 55884 | EPI_ISL_6811664 |
| 7. | Delta | 18979 | 1.126e+05 | 98% | 0.0 | 99.68 | 55947 | EPI_ISL_6864866 |
| 8. | Gamma | 19038 | 1.093e+05 | 96% | 0.0 | 99.33 | 56110 | EPI_ISL_6864534 |

#### S1.2.2 Spike protein sequence

| Ranking | SARS-CoV-2 strain | Max score | Total score | Query cover | E value | Percentage Identity | Accession length | GISAID/NCBI Accession number |
|---|---|---|---|---|---|---|---|---|
| 1. | Delta | 2556 | 14331 | 100% | 0.0 | 96.71 | 55947 | EPI_ISL_6864866 |
| 2. | Alpha | 2536 | 14061 | 100% | 0.0 | 97.01 | 55890 | EPI_ISL_6865096 |
| 3. | Mu | 2534 | 14051 | 100% | 0.0 | 97.02 | 55884 | EPI_ISL_6811664 |
| 4. | WT with D614G mutation (B.1) | 2533 | 14031 | 100% | 0.0 | 97.02 | 56235 | EPI_ISL_6832722 |
| 5. | Wild type (WT) | 2531 | 14021 | 100% | 0.0 | 96.94 | 56040 | NC_045512.2 |
| 6. | Gamma | 2522 | 13949 | 100% | 0.0 | 96.71 | 56110 | EPI_ISL_6864534 |
| 7. | Beta | 2522 | 13934 | 100% | 0.0 | 96.71 | 55693 | EPI_ISL_6838995 |
| 8. | Lambda | 2496 | 13804 | 100% | 0.0 | 96.00 | 55954 | EPI_ISL_6694892 |

#### S1.2.3 Receptor binding motif (RBM) protein sequence

| Ranking | SARS-CoV-2 strain | Max score | Total score | Query cover | E value | Percentage Identity | Accession length | GISAID/NCBI Accession number |
|---|---|---|---|---|---|---|---|---|
| 1. | Alpha | 148 | 813 | 100% | 2e-39 | 90.41 | 55890 | EPI_ISL_6865096 |
| 2. | Mu | 148 | 805 | 100% | 2e-39 | 90.41 | 55884 | EPI_ISL_6811664 |
| 3. | Beta | 148 | 805 | 100% | 2e-39 | 90.41 | 55693 | EPI_ISL_6838995 |
| 4. | Gamma | 148 | 802 | 99% | 2e-39 | 90.41 | 56110 | EPI_ISL_6864534 |
| 5. | Delta | 147 | 803 | 100% | 4e-39 | 90.41 | 55947 | EPI_ISL_6864866 |
| 6. | Wild type (WT) | 145 | 793 | 100% | 2e-38 | 89.04 | 56040 | NC_045512.2 |
| 7. | WT with D614G mutation (B.1) | 145 | 793 | 100% | 2e-38 | 89.04 | 56235 | EPI_ISL_6832722 |
| 8. | Lambda | 141 | 785 | 100% | 3e-37 | 87.67 | 55954 | EPI_ISL_6694892 |

*SARS-CoV-2/human/BEL/rega-20174/2021, complete genome, GenBank: OL672836.1, 29684 bp
